# Serious harms of the COVID-19 vaccines: a systematic review

**DOI:** 10.1101/2022.12.06.22283145

**Authors:** Peter C. Gøtzsche, Maryanne Demasi

## Abstract

**BACKGROUND:** Serious and severe harms of the COVID-19 vaccines have been downplayed or deliberately excluded by the study sponsors in high impact medical journals.

**METHODS:** Systematic review of papers with data on serious adverse events (SAEs) associated with a COVID-19 vaccine.

**RESULTS:** We included 18 systematic reviews, 14 randomised trials, and 34 other studies with a control group. Most studies were of poor quality. A systematic review of regulatory data on the two pivotal trials of the mRNA vaccines found significantly more SAEs of special interest with the vaccines compared to placebo, and the excess risk was considerably larger than the benefit, the risk of hospitalisation. The adenovirus vector vaccines increased the risk of venous thrombosis and thrombocytopenia, and the mRNA-based vaccines increased the risk of myocarditis, with a mortality of about 1-2 per 200 cases. We found evidence of serious neurological harms, including Bell’s palsy, Guillain-Barré syndrome, myasthenic disorder and stroke, which are likely due to an autoimmune reaction. Severe harms, i.e. those that prevent daily activities, were underreported in the randomised trials. These harms were very common in studies of booster doses after a full vaccination and in a study of vaccination of previously infected people.

**CONCLUSIONS:** Further randomised trials are needed. Authorities have recommended populationwide COVID-19 vaccination and booster doses. They do not consider that the balance between benefits and harms becomes negative in low-risk groups such as children and people who have already recovered from COVID-19 infection.

## Introduction

Vaccines to prevent SARS-CoV-2 infection were developed to curb the COVID-19 pandemic. Major drug regulators such as the US Food and Drug Administration (FDA) and the European Medicines Agency (EMA), authorised the first COVID-19 vaccines under emergency or conditional use in December 2020 through accelerated pathways,^1,2^ which provides for a lower level of evidence for effectiveness than standard product approvals.^3^ Authorities stated that the vaccines were highly effective at preventing infection and severe disease, and, in total, only one severe case of COVID-19 occurred in the vaccine groups compared with 49 in the control groups in the three pivotal trials from Pfizer, Moderna and AstraZeneca.^4-6^ Governments commenced population-wide vaccination campaigns immediately, prior to the completion of any of the conventional phases of clinical trials or any medium or long-term harms could be elucidated.

Serious concerns have been raised about the reliability of clinical trial data, partly because the pharmaceutical industry has a history of falsifying data and deliberately hiding harms.^7^ Also, neither the vaccine manufacturers, nor the drug regulators have allowed independent researchers access to the raw trial data of the COVID-19 vaccines.^8^ Transparency advocates have sued the FDA for access and a court ordered the agency to release regulatory documents.^9^

Previously, we documented that serious harms have been excluded from the published trial reports of the COVID-19 vaccines.^10,11^ However, data from other types of research, mainly pharmacovigilance studies, have associated thrombosis, myocarditis and the Guillain-Barré syndrome with COVID-19 vaccination.^12^

We performed a systematic review of the published studies on all types of COVID-19 vaccines to analyse the risk of serious harms.

## Methods

We carried out a systematic review of systematic reviews and observational studies that included data on serious adverse events (SAEs) associated with a COVID-19 vaccine. According to EMA, an SAE is an adverse event that results in death, is life-threatening, requires hospitalisation or prolongation of existing hospitalisation, results in persistent or significant disability or incapacity, or is a birth defect.

In clinical trials, the severity of adverse events is often classified into mild, moderate, and severe where severe means preventing usual activity.

We noted in our protocol that we might limit the inclusion of reviews and studies according to methodological rigour or number of patients, if the workload became excessive. This was the case, and we therefore excluded studies that addressed special groups of people, e.g. patients with inflammatory bowel disease and pregnant women; studies based on questionnaires; studies that did not have a comparator group; and randomised trials and comparative cohort studies that had less than 1000 participants. We also needed to abandon our aim of reviewing adverse events lasting at least one year, as our searches did not provide such data.

### Search strategy and selection of studies

We searched PubMed on 4 April 2022 with this strategy: (COVID-19 OR SARS-CoV-2) AND (vaccin*) AND (safety OR adverse event* OR harm*).

One researcher (MD) screened the search results by title and abstract and excluded articles that clearly did not fulfil our inclusion criteria. Any records where there was doubt were examined by both researchers. Next, we examined the full reports for possible inclusion independently, resolving disagreements by discussion.

### Data management and data extraction

We used Zotero to manage the search results and MS Excel and Word to handle the extracted data. One researcher extracted data, and doubts were resolved by discussion.

We described the risks for adverse events and focused on bias and confounders in the studies. As we expected significant heterogeneity in the way the studies were carried out and reported, we aimed primarily at producing a narrative systematic review, which could be useful for decision making and for planning research.

### Statistical methods

We did a meta-analysis using a random effects model with the Comprehensive Meta Analysis software for serious adverse events in children.

## Results

Our search yielded 4,637 records. We initially excluded 4,074 obviously irrelevant records. After examining the remaining records, we excluded another 479 records: 242 cohort studies without an adequate control group, 36 comparative studies with less than 1000 participants, five reports with data included in other papers, a study of a typhoid vaccine, a small study with a meningococcal vaccine in the control group, 126 reports of single cases, 61 reports of multiple cases, five reports with no cases, and two studies based on questionnaires.

We also excluded 26 of the 42 systematic reviews we found: 13 did not look for serious adverse events or reported that there were none (including a review about “safety and efficacy” of the vaccines with over 100,000 patients from randomised trials);^13^ three were in pregnancy; one from Wuhan in China which did not report SAEs by treatment group;^14^ two were about inflammatory bowel disease; one about eye diseases; two were not about COVID-19 vaccines; one was a protocol; one was an autopsy study that established a causal relationship in 15 of 38 deaths;^15^ and one from Hong Kong was unreliable, as it combined data from trials with those from observational studies and concluded that the 95% confidence intervals did not indicate a relationship between the vaccines and SAEs, which was incorrect as several confidence intervals excluded the possibility of no relationship.^16^

We included 17 systematic reviews,^17-33^ 14 randomised trials,^34-47^ and 31 other studies with a control group.^48-78^ Four of these were not identified in our search. A systematic review^17^ and two registry studies^53,78^ were published after the cut off for our search, and a self-controlled case series study was provided by a colleague.^66^

### Serious adverse events in general

The most methodologically rigorous, reliable, and relevant research paper we retrieved was a systematic review conducted by researchers from USA, Spain, and Australia of regulatory data on the two pivotal randomised trials of the mRNA vaccines, one from Pfizer and one from Moderna.^17^

The review analysed SAEs in general and SAEs of special interest (AESI) according to two lists for the Brighton Collaboration criteria adopted by the WHO. The trials were expected to follow participants for two years. However, within weeks of the FDA emergency use authorisation, the sponsors began to unblind the participants and offered the vaccine to those in the placebo group.^17^ Therefore, the authors used the interim datasets that were the basis for the emergency authorisation, covering about four months after the trials commenced.

The authors included SAEs results tables from the websites of the FDA and Health Canada. Based on blinded tables, two clinicians judged independently whether an SAE was also an AESI. To account for multiple SAEs occurring in the same patient, a standard adjustment was used to widen the confidence intervals.

For SAEs, the risk difference for the two vaccines was 13.2 per 10,000 vaccinated people (95% confidence interval -3.2 to 29.6) and the risk ratio was 1.16 (0.97 to 1.39).

For SAEs of special interest, the risk difference and the risk ratio were significantly increased, 12.5 (2.1 to 22.9) and 1.43 (1.07 to 1.92), respectively. The largest excess risk occurred amongst the Brighton category of coagulation disorders (36 patients in the vaccine groups and 23 in the placebo groups). Only 6 vs 6 patients developed myocarditis/pericarditis.

Even though the researchers blinded their classifications, critics have claimed that they should have excluded some events and included others. Subsequently, the researchers redid their analyses based on the criticism, and it actually rendered the results slightly worse for the vaccines (Peter Doshi, personal communication).

The SAEs in the Moderna trial were misleading. For reasons not documented in the trial protocol, Moderna included efficacy outcomes in its SAEs tabulations, while Pfizer excluded them. Thus, COVID-19 complications were counted as SAEs and these were more common in the placebo group, and therefore, skewed the results.

Pfizer’s vaccine increased SAEs significantly, risk difference 18.0 per 10,000 (1.2 to 34.9) and risk ratio 1.36 (1.02 to 1.83). In contrast, FDA concluded that SAEs were “balanced between treatment groups.” This discrepancy may in part be explained by the fact that FDA analysed participants experiencing one or more SAEs because they had access to individual participant data, whereas the researchers did not, and therefore analysed total SAEs. Hence, FDA’s analysis did not reflect the observed excess of multiple SAEs in the vaccine group. More importantly, FDA used a different analysis population, which resulted in 126 vs 111 participants with SAEs whereas the researchers found 127 vs 93, also using FDA data.

Systematic reviews of mainly published trials were of poor quality and flawed. One from India included both randomised and non-randomised studies and did not find an increase in SAEs: 0.7% in people receiving the AstraZeneca vaccine and 0.8% in the control groups.^18^ The authors stated that their search strategy, ‘‘(COVID-19 Vaccine)” retrieved 196 records, but when we repeated it for the same time period, we retrieved 3,371 records. Some of the data were also erroneous. In a table, the authors stated that there were only 4 SAEs in Pfizer’s pivotal trial,^5^ but in fact, there were 126 vs 111, which they described as 126 vs 11, in the text.

A Chinese review did not find an increase in the risk of SAEs, risk ratio 0.94 (0.71 to 1.25), and the vector-based vaccines decreased the risk of SAEs, risk ratio 0.79 (0.63 to 0.99).^19^

Another Chinese review only presented data in a supplement, divided by organ class, with no statistical estimates.^20^

A review from Indonesia presented no summary data on SAEs.^21^

A review from Canada, of 25 randomised trials and 105,527 patients, only mentioned three anaphylactic shocks on the vaccine and one on placebo.^22^

In a follow-up of Pfizer’s trial, 24 of the 32 authors were from Pfizer.^34^ Even though the additional data contributed to the full approval of the vaccine in the United States, there were no numerical data on SAEs in the trial report in *New England Journal of Medicine*, which just noted that no new SAEs “were considered by the investigators to be related to BNT162b2” and that “No new safety signals relative to the previous report were observed during the longer follow-up period.” This was highly misleading. The journal article specified that safety would be evaluated through 6 months after the second dose, but what was published in a supplement on a website was in violation of Pfizer’s own protocol and the study report. The supplement only showed data reported up to one month after the second vaccine dose. Thus, Pfizer had omitted five months of safety data. Deliberately hiding harms data could be considered fraud.

In a trial of Janssen’s vaccine, 19 of the 20 authors were from Janssen.^35^ SAEs occurred in 223 of 21,898 patients on vaccine vs 265 of 21,890 patients on placebo, and 19 vs 2 patients had SAEs considered by the investigator to be related to vaccination. The authors noted the following imbalances in adverse events occurring within 28 days after vaccination: tinnitus (15 vs 4), urticaria (13 vs 6), convulsion (9 vs 4), pulmonary embolism (10 vs 5), and deep vein thrombosis (11 vs 3). We calculated that the vaccine reduced total mortality, 28 vs 55 deaths, risk ratio 0.51 (0.32 to 0.80), and COVID-19 related mortality, 5 vs 22 deaths, risk ratio 0.23 (0.09 to 0.60). The authors found the same but used person-years as denominators.

In a trial of AstraZeneca’s vaccine, 101 of 21,587 patients (0.5%) vs 53 of 10,792 (0.5 %) had an SAE within 28 days after a vaccine dose.^36^ The paper specified that SAEs would be recorded from “the time of signed informed consent through day 730.” However, there were no data for SAEs beyond 28 days. As it is implausible that no one of 32,379 patients would be admitted to hospital (which is a SAE), for two years, many SAEs must be missing, not only from the trial report but also in its supplementary data. This trial was published in *New England Journal of Medicine*. There were 7 vs 7 deaths in the whole trial period.

A trial in India of ZyCoV-D, a DNA-based vaccine, was also highly problematic. It randomised 27,703 patients, either aged 12-17 years or 60 years and older.^37^ A supplement reported one SAE in the vaccine group and none in the placebo group among the elderly and one vs two in “comorbid subjects.” The main text was totally different, with no division as per randomised group. It described 15 SAEs, but seven of these were merely being COVID-19 positive, which is not an SAE and furthermore belongs to the reporting of the benefits, not the harms. There was one death in each group. This paper, which was difficult to interpret, was published in *The Lancet*.

In a UK trial of a recombinant nanoparticle vaccine from Novavax (NVX-CoV2373), published in *New England Journal of Medicine*, there were 41 patients with SAEs of 7,569 in the vaccine group and 41 of 7,570 in the placebo group in one table in a supplement, but in another supplement table, the numbers were 44 vs 44 SAEs.^38^

In a US-Mexican trial, also of the NVX-CoV2373 vaccine, a supplement showed that 228 of 19,729 patients (1.2%) had an SAE in the vaccine group and 128 of 9,853 (1.3%) in the placebo group.^39^ Treatment-emergent systemic adverse events grade 4 within 7 days (which are life-threatening) were more common in the vaccine group, 17 vs 5 after first dose and 21 vs 5 after second dose. There was no mention of grade 4 events in the main text. The trial was published in *New England Journal of Medicine*.

An Indonesian trial of inactivated SARS-CoV-2 whole virion vaccine from Sinovac randomised 1620 people:^40^ “there were nine serious adverse events (SAEs) that occurred in all subjects with a classification not related to vaccine products (five SAEs).”

It was not possible to interpret the text in this paper, published in *Vaccine*.

A Taiwanese trial of a recombinant protein subunit vaccine (MVC-COV1901) provided no data in the article, which only stated that “No serious adverse events were considered related to the study intervention.”^41^ However, in a supplement, 18 of 3295 patients (0.6%) had an SAE on the vaccine and 1 of 549 (0.2%) on placebo. Unsolicited adverse events grade 3 or above occurred in 93 vs 11 patients. Grade 3 was not defined, but it is commonly defined as being serious and interfering with a person’s ability to do basic things like eat or get dressed. The trial was published in *Lancet Respiratory Medicine*.

In an Indian trial of the AstraZeneca vaccine, 12 of 900 patients had an SAE on the vaccine and 2 of 300 on placebo.^42^

A US register study of nursing home residents reported lower 7-day mortality after first vaccination than among unvaccinated people, risk ratio 0.34 (0.22 to 0.54) but no difference in hospitalisations, risk ratio 0.95 (0.72 to 1.24).^48^ These results are not reliable, as the researchers adjusted for 11 confounders (see Discussion).

### Thromboses

Most systematic reviews were of poor quality. A Canadian review of randomised trials described 37 blood clots in the Results section on the AstraZeneca vaccine, but they did not come from the trials but from 17 million vaccinated people, which is 0.2 cases per 100,000.^22^

A systematic review of non-randomised studies from South Korea identified 664 patients who developed vaccine-associated thrombosis with thrombocytopenia after an adenovirus vector vaccine.^23^ The mean age was 46 years, 70% were females, 91% had antibodies against platelet factor 4, and 32% died. The pooled incidence of venous thrombosis after Astra-Zeneca’s vaccine was 28 (12-52) per 100,000 doses, or 130 higher than in the Canadian study. The pooled incidence rate of cerebral venous thrombosis after the AstraZeneca vaccine was much higher than the background rate, 23 vs 0.9 per 100,000 person-years.

A systematic review from USA, mainly of case reports, identified 144 patients with thromboembolic events after the AstraZeneca vaccine.^24^ The mean age rage was 21 to 68 years, 65% were females, and 75% had thrombocytopenia. Mean time for onset of symptoms was 8 days; 50% died. The denominators vary, which makes it difficult to interpret the review.

A systematic review from Pakistan of case reports identified 80 patients with cerebral venous sinus thrombosis after vaccination.^25^ In 83% of cases, the patients had received an adenovirus vector vaccine. The mean age was 43 years, 74% were females, and 56% had antibodies against platelet factor 4. Mean time for onset of symptoms was 11 days; 39% died.

Another systematic review from Pakistan of case reports included 65 patients with thrombosis with thrombocytopenia after vaccination.^26^ In 92% of cases, the patients had received an adenovirus vector vaccine. The mean age was 54 years, 79% were females, and 82% had antibodies against platelet factor 4. Some numbers were wrong, e.g. 36 of 51 females survived and 15 died but the percentages were 80% and 62.5% respectively. Mean time for onset of symptoms was 9 days; 37% died.

A systematic review from Qatar included mainly case reports but also five observational studies and one “multinational study.”^27^ The authors in- and excluded studies along the way. We were unable to extract any meaningful data from this study.

In a self-controlled case series study of hospital admissions and deaths based on UK register data, the risk of thrombocytopenia was increased after the AstraZeneca vaccine, incidence rate ratio 1.33 (1.19 to 1.47) and after a SARS-CoV-2 infection, 5.27 (4.34 to 6.40).^49^ The risk was also increased for venous thromboembolism, 1.10 (1.02 to 1.18) and 13.86 (12.76 to 15.05), respectively, and for cerebral venous sinus thrombosis, 4.01 (2.08 to 7.71) and 13.43 (1.99 to 90.59), respectively, where the risk was also increased for Pfizer’s vaccine, 3.58 (1.39 to 9.27).

The risk of arterial thromboembolism was increased for Pfizer’s vaccine and after a SARS-CoV-2 infection, 1.06 (1.01 to 1.10) and 2.02 (1.82 to 2.24), respectively. The risk was also increased for ischaemic stroke, 1.12 (1.04 to 1.20) and 2.00 (1.70 to 2.35) after an infection, respectively, and for other rare arterial thrombotic events after the AstraZeneca vaccine, 1.21 (1.02 to 1.43). Censoring the data to the time before concerns about thrombosis were raised made no difference, and the incidence of coeliac disease, which was a negative control outcome, did not change.

A study from Scotland using a national cohort also found an increased risk of thrombocytopenia after the AstraZeneca vaccine, adjusted rate ratio 5.77 (2.41 to 13.83), which was confirmed in a self-controlled case series analysis, risk ratio 1.98 (1.29 to 3.02).^50^

Indian researchers used the VigiBase for disproportionality analyses, but their methods were doubtful, and they did not explain what the COVID vaccines were compared with.^51^ They noted that, “based on IC_025_ values, acute myocardial infarction, cardiac arrest, and circulatory collapse were associated with the vaccines used in the age group > 75 years.”

A UK study used registry data from 8 December 2020 to 18 March 2021, in which period 21 of 46 million had their first vaccination.^52^ The researchers adjusted their estimates for a total of 30 confounders, and they used lower limb fracture as a control condition unlikely to be affected by vaccination. However, there were significantly fewer fractures after vaccination. In the Discussion, the authors mention six limitations but do not discuss fractures and avoid mentioning that their data on fractures render their data unreliable. For example, the authors reported a hugely protective effect of the AstraZeneca vaccine against venous thrombosis in the elderly (at least 70 years of age), hazard ratio 0.58 (0.53 to 0.63), whereas other research shows that this vaccine causes thrombosis. Data on all-cause mortality were also implausible, e.g. a hazard ratio of 0.19 (0.19 to 0.20) after the Pfizer vaccine. It is hard to imagine that a COVID-vaccine could reduce total mortality by 80% in elderly people, as they die from so much else. The study was published in *PLoS Medicine*.

In a European-US register study, the researchers estimated incidence rate ratios in adults after propensity scores matching and calibration using 92 negative control outcomes.^53^ The statistical methods were highly complex and involved nine confounders. Compared with Pfizer’s vaccine, the AstraZeneca vaccine increased the risk of thrombocytopenia, rate ratio 1.33 (1.18 to 1.50), risk difference 8.21 (3.59 to 12.82) per 100,000 recipients. The paper is difficult to interpret because there is an enormous amount of data on various types of thromboses; the data from country to country are not consistent; there were systematic errors, especially in the US Open Claims database; and immunisation practices were different. There was no increase in myocardial infarction.

A registry study of Danish frontline workers included data from 27 Dec 2020 to 13 April 2021.^54^ Even though people were their own controls, the outcomes were adjusted for 10 confounders. The AstraZeneca vaccine increased the risk of deep vein thrombosis, risk difference 8.4 (0.2 to 16.5) per 100,000 vaccinations. The Pfizer vaccine but not the AstraZeneca vaccine reduced the mortality risk; the risk difference was -4.2 (−8.2 to -0.1) and -1.6 (−7.2 to 4.0), respectively. These results are the opposite of those from the randomised trials, where the AstraZeneca vaccine lowered mortality, risk ratio 0.37 (0.19 to 0.70), but the Pfizer vaccine didn’t, risk ratio 1.03 (0.63 to 1.71).^79^ This suggests that when analyses are adjusted for many confounders, this may negate the advantage of using people as their own controls.

Italian researchers used the EudraVigilance European database to compare the vaccines from Astra-Zeneca, Janssen and Pfizer for cardiovascular, neurological, and pulmonary events.^55^ The paper is uninterpretable. They mistakenly talk about severe adverse events, abbreviated as SAEs, when the events are serious, which is worse than just being severe; the issue with confounders didn’t even appear in their 10,856 word article; the age was unknown in over half of the people vaccinated; and they present-ed large hazard ratios with no confidence intervals.

In a similar study by some of the same authors, the risk ratios for cerebral vein thrombosis, splanchnic vein thrombosis, thrombocytopenia, and other bleeding events in people at least 65 years of age were 2-7 times higher for the AstraZeneca vaccine than for the Pfizer vaccine, with narrow confidence intervals.^56^ The data used were those added to data bank until 16 April 2021, before concern was raised about the AstraZeneca vaccine causing blood clots.

The authors noted that while EMA reported only one SAE per million vaccine doses related to blood clots and thrombocytopenia, they found 151 and 36, respectively, for the two vaccines, with 13 and 4 deaths possibly related to this. They also reported that SAEs in the categories “nervous system disorders”, “gastrointestinal disorders” and “musculo-skeletal and connective tissue disorders” occurred 9 times more often with the AstraZeneca vaccine than with the Pfizer vaccine but listed no confidence intervals. Yet again, they called serious events severe events.

In a French registry study of people at least 75 years old where the patients were their own controls, the researchers wrote that in the first two weeks after each dose of Pfizer’s vaccine, “no significant increased risk was found for any outcome.”^57^ They actually found a *decreased* risk after the first dose for ischaemic stroke, relative incidence 0.90 (0.84 to 0.98) and for pulmonary embolism, 0.85 (0.75 to 0.96).

A registry study with US and Indian authors was seriously misleading.^58^ The title was declarative: “Cerebral venous sinus thrombosis is not significantly linked to COVID-19 vaccines or non-COVID vaccines in a large multi-state health system,” but the study was vastly underpowered and unable to detect anything. There were only 3 cases after Pfizer’s vaccine and none after Moderna’s vaccine. The abstract was also misleading. There were no numerical data, only a mention of “not significantly associated.”

Italian researchers used data on cerebral vein thrombosis reported to the EudraVigilance database during the first six months of 2021.^59^ The reporting rate per million people receiving their first dose of vaccine was 21.6 (20.2 to 23.1) for AstraZeneca, 11.5 (9.6 to 13.7) for Janssen, 5.6 (4.7 to 6.6) for Moderna and 1.9 (1.7 to 2.1) for Pfizer. Cerebral vein thrombosis occurred alongside thrombocytopenia with all four vaccines, and the observed to expected ratio was significantly increased for all four vaccines, also using the highest estimated background incidence. Two limitations of the study are that the use of the vaccines in various age groups was not the same throughout Europe and that half of the observation period was after EMA had raised concern about possible blood clots caused by the adenovirus vector vaccines.^80^

A study from India reported on 89 patients with acute coronary syndrome, 37 of whom had a prior vaccination history.^60^ It is not possible to conclude anything about possible vaccine harms based on this paper.

### Myocarditis and pericarditis

A systematic review from India included 2184 patients with myocarditis.^28^ The mean age was 26 years, 73% were males, and 99% had received an mRNA-based vaccine. Mean time for onset of symptoms was 4 days. The paper is difficult to comprehend, e.g. 1339 patients had definite, probable or possible myocarditis but there were 845 more patients with myocarditis, and the percentage of patients admitted to the intensive care unit is derived from a denominator of only 1169. Six patients died among 1317 for which data were available, a rate of one per 200.

A systematic review from Singapore included published articles based on five vaccine safety surveillance databases and 52 case reports totalling 200 cases of possible COVID-19 vaccine-related myocarditis.^29^ The authors tried to cover too much ground in one article, which makes it difficult to read, and what they found was not new and has been better described by other authors.

A systematic review with European authors included 129 cases,^30^ but cannot be used for a risk assessment.

A systematic review of myocarditis after an mRNA vaccine included data from 69 patients based on case reports and case series.^31^ The mean age was 21 years, 93% were males, and 89% developed symptoms after the second dose. Patients were admitted to hospital a median of three days post-vaccination.

A systematic review from China of children and adolescents included both randomised trials, observational studies, and case reports.^32^ The authors “summarized the basic information of 27 cases from included studies,” which did not allow a risk assessment.

In a self-controlled case series study of hospital admissions and deaths based on UK registry data, the AstraZeneca vaccine and the mRNA vaccines increased the risk of myocarditis, with incidence rate ratios between 1.33 and 1.72, which were lower than the risk after a SARS-CoV-2 infection, 11.14.^61^

The authors confirmed their results in a similar study, which found decreased risks of cardiac arrhythmia, apart from an increase after the second dose of Moderna’s vaccine, incidence rate ratio 1.93 (1.25 to 2.96 at 1-7 □ days).^62^ There was no increased risk of encephalitis, meningitis and myelitis after the vaccines from AstraZeneca and Pfizer, 1.07 (0.87 to 1.31) and 1.14 (0.86 to 1.51), respectively, whereas infection increased the risk, 2.07 (1.78 to 4.11).

A French disproportionality study of myocarditis and pericarditis after an mRNA vaccine reported to VigiBase, the WHO’s pharmacovigilance database, included data till June 2021.^63^ Compared with older patients, myocarditis was much more commonly reported in young people; the reporting odds ratio (ROR) was 22.3 (19.2 to 25.9) for adolescents and 6.6 (5.9 to 7.5) for 18–29 years old. Myocarditis was much more common in males, ROR 9.4 (8.3 to 10.6). Median time to onset was 3 days for myocarditis and 8 for pericarditis; 21% of the cases with inflammatory heart reactions were life-threatening, and 1% died (22 of 2277 cases). The estimated rate of myocarditis was 3.6 (3.3 to 3.9) per 100,000 fully vaccinated persons in USA, and 7.8 (6.9 to 8.9) in young adults.

A US study using the Vaccine Adverse Event Reporting System (VAERS) found that patients with myocarditis after an mRNA vaccine reported between December 2020 and August 2021 had a median age of 21 years and that 82% were males.^64^ The incidence in young males was over 10 times higher than in middle-aged males, and 82% of cases occurred after the second vaccination. The reporting rates in young adults were 30 times higher than the expected background rate. Glucocorticoids were used in 12% of the patients, but the most common treatment was nonsteroidal anti-inflammatory drugs, used in 87% of the patients. This is surprising because these drugs, despite their name, have no anti-inflammatory properties^81-83^ and increase the risk of heart attacks and death.^7^

Another US VAERS study, with US and Chinese authors, came to different results even though it used the same observation period.^65^ The adverse event rate in adolescents was three times higher than in the former study, which cannot be explained by inclusion also of pericarditis and by having no 7-day limit for reporting. Most cases occur within the first couple of days and myocarditis is diagnosed about 10 times as often as pericarditis.^65^ The risk was greater for Pfizer’s vaccine, ROR 5.4 (4.1 to 7.0) than for Moderna’s, ROR 2.9 (2.2 to 3.8), but, as the authors noted, only Pfizer’s vaccine was approved for use in minors where the risk is greatest, and the risks were similar in other age groups.

The authors wrote that Janssen’s vaccine “was not associated with signals of myocarditis/pericarditis.” This statement is extremely misleading. First, even though few people received this vaccine, the estimate was very close to being statistically significant, ROR 1.39 (0.99 to 1.97), which is surely a signal. Second, Janssen’s vaccine was only approved for adults. Third, the authors wrote that the incidence rate was higher after the mRNA vaccines than after viral vector vaccines, but they reported that these rates were 5.98 (5.73 to 6.25) vs 5.64 (4.46 to 7.04) per million, which are similar rates, and the confidence interval for Janssen’s vaccine includes the whole confidence interval for the mRNA vaccines. We looked up if the authors had a conflict of interest related to Janssen, but they declared they had none.

### Inflammatory neuropathies

In the randomised trials, there were seven cases of Bell’s palsy among people receiving an mRNA vaccine versus one among placebo recipients (P = 0.07), and the incidence rate was 3.5-7 times higher than the background rate.^69^

This signal was also found in a self-controlled case series study of hospital admissions based on UK register data.^66^ There was an increased risk of Bell’s palsy, incidence rate ratio 1.29 (1.08 to 1.56), Guillain-Barré syndrome, 2.90 (2.15 to 3.92), and myasthenic disorder 1.57 (1.07 to 2.30) with the AstraZeneca vaccine. Pfizer’s vaccine increased the risk of haemorrhagic stroke, 1.38 (1.12 to 1.71). The risk of neurological outcomes was also increased after infection with SARS-CoV-2. There were 4 excess cases of Guillain-Barré syndrome per million people receiving the AstraZeneca vaccine and 15 excess cases after an infection.

No such signals were found in a study using data from primary care records in the UK and Spain, not even in a series of self-controlled cases of Bell’s palsy.^67^ The risks for Bell’s palsy, Guillain-Barré syndrome, and encephalomyelitis were lower than expected background rates or about the same for the vector based and mRNA vaccines.

A case-control study from Israel with 37 cases of facial nerve palsy did not find an association to Pfizer’s vaccine, odds ratio 0.84 (0.37 to 1.90).^68^

An Israeli register study of Pfizer’s vaccine compared the rates of Bell’s palsy with background rates.^69^ The standardised incidence ratio after the first dose was 1.36 (1.14 to 1.61). This is a weak signal in a study with a historical control. Expected cases cannot be determined with sufficient precision and they vary over time. The signal was even weaker after the second dose, 1.16 (0.99 to 1.36). In elderly females where the strongest association was observed in this study, the excess risk of Bell’s palsy was estimated to be 5 cases per 100,000 vaccinees.

Another Israeli register study matched vaccinated with unvaccinated people for seven factors and adjusted for socioeconomic status for which matching was poor.^70^ Pfizer’s vaccine did not increase the occurrence of Bell’s palsy, risk ratio 0.96 (0.54 to 1.70) or Guillain-Barré syndrome (1 vs 0 cases), whereas there were more cases of numbness or tingling, risk ratio 1.22 (1.08 to 1.37).

Using VigiBase for disproportionality analyses, Swiss researchers found *lower* risks for COVID-19 vaccines than for other viral vaccines for neuralgic amyotrophy, ROR 0.23 (0.17 to 0.30) vs 0.12 (0.09 to 0.16), and for Guillain-Barré syndrome, ROR 0.15 (0.13 to 0.16) vs 0.06 (0.05 to 0.06).^71^ In contrast, Bell’s palsy was more frequently reported with COVID-19 vaccines, ROR 1.12 (1.07 to 1.17). Indian researchers also used VigiBase for disproportionality analyses, but their methods and conclusions were doubtful.^72^ They referred to IC_025_ values without explaining what it meant and did not state what the COVID vaccines were compared with. They listed 52 neurological diagnoses, which they “considered to be associated with the administration of the vaccine.”

A systematic review from Kuwait and Egypt was also problematic, e.g. there was no reproducible search strategy,^33^ which is essential for systematic reviews. The authors reported on 32 cases of CNS demyelination following various COVID-19 vaccines.

A study based on 555 reports in VAERS of hearing loss did not find an increase in risk, compared to the background rate.^73^

### Serious adverse events in people with previous infection

In an Israeli study, Pfizer’s vaccine was given to 78 people with a previous COVID-19 infection and to 177 matched controls.^74^ Some numbers and percentages are erroneous. Emergency department visit or hospitalisation was required for 5 (6%) vs 1 patients (0.6%). Even though the authors showed in a table that this difference was statistically significant (P = 0.01), they concluded that the vaccine was safe in people with previous infection. This is not correct. Hospitalisation is a serious harm, and harms occurred ten times as often if the patients had been infected earlier, suggesting that those with acquired immunity are at higher risk of experiencing SAEs post vaccination.

### Serious adverse events after a booster dose

In a US study, 305 people previously vaccinated with two doses of 100 μg of Pfizer’s vaccine received a third, booster dose and were compared with the second dose of the vaccine in 584 historical controls and with a 50 μg booster in separate studies.^75^

The 100 μg booster caused more local and systemic adverse reactions than the second 100 μg vaccine dose and the 50 μg booster. A supplement showed that there was a large difference for moderate or severe solicited systemic adverse reactions; 59% experienced this on the 100 μg booster vs 39% on the 50 μg booster (P = 0.000,05, our calculation). There was no such difference between the 100 μg booster and the 100 μg second dose, 59% vs 54% (P = 0.12).

There were two serious adverse events (not six, as the authors claim, as the other four were asymptomatic infections with positive tests) but no information about which groups they came from.

In another US study, the patients used v-safe, a voluntary, smartphone-based safety surveillance system developed by the Centers for Disease Control and Prevention to provide information on adverse reactions after vaccination.^76^ The occurrence of adverse reactions was very similar for dose three and dose two (99.7% of the doses were mRNA vaccines). There were many severe adverse events: 28% were unable to perform normal daily activities after the booster; 11% were unable to work or attend school; 0.2% had an emergency visit; and 0.1% were hospitalised.

In a US study of Pfizer’s vaccine, conducted by Pfizer, patients were randomised to receive a third dose or placebo.^43^ After a third dose, 16 of 5,055 patients had an SAE on the vaccine and 24 of 5,020 on placebo. The study was published in *New England Journal of Medicine*, and 24 of the 32 authors were from Pfizer or hired by Pfizer.

In contrast, in a self-controlled case series study of hospital admissions based on UK registry data, the risk of myocarditis was increased after a booster dose of Pfizer’s vaccine, incidence rate ratio 1.72 (1.33 to 2.22).^61^

A UK study of 2,878 people was uninterpretable, as they were randomised to 12 different groups including a meningococcal vaccine and as there were only 24 SAEs.^44^

### Serious adverse events in children

We found three randomised trials with data on SAEs in children. In all cases, the data were hidden in supplements to the article. In two trials of mRNA vaccines, 6 of 2486 vs 2 of 1240 children 12-17 years of age and 4 of 1131 vs 1 of 1129 children 12-15 years of age had SAEs, respectively.^45,46^ In a trial of a Chinese attenuated virus, the term SAEs was not used but the numbers for grade 3 reactions were 1 of 251 vs 0 of 84 in children 6-12 years of age.^47^ The pooled risk ratio for these three trials was 1.90 (0.57 to 6.29, P = 0.29, I^2^ = 0).

### Other issues

Appendicitis has been suggested as a possible adverse because of a numerical increase in a vaccine trial.^53f^ A US study reviewed cases of appendicitis reported to VigiBase and found 358 cases compared to 329 expected cases.^77^ We explored this and found a Danish registry study that reported an adjusted risk ratio of 0.93 (0.79 to 1.11) after the first dose and 0.99 (0.84 to 1.18) after the second dose of the suspected agent, an mRNA vaccine.^78^

## Discussion

Our systematic review demonstrates the difficulty of determining vaccine related SAEs in published trial data. Theoretically, systematic reviews of randomised trials should be the most reliable source of evidence, but serious harms are vastly underreported, if reported at all, in published drug trials.^7^

The underreporting seems to be particularly pronounced in vaccine trials.^10,11,84^ For the COVID-19 vaccines, there is the additional problem that, within weeks of the vaccines receiving an emergency use authorisation, when far too little time had elapsed to identify late occurring or diagnosed harms, the unblinding of trials commenced and placebo recipients were offered the vaccine.^85^

The safety of vaccines is important because they are preventive, but editors of our most prestigious journals allowed the data on serious harms to be relegated to supplements, which few readers will access, particularly if they read the paper version.

Severe harms – which are defined as those preventing usual activity – have also been vastly underreported in the published trial reports. Pfizer’s pivotal trial report, published in *New England Journal of Medicine*, was highly misleading.^5^ It mentioned only serious adverse events considered related to the vaccine: four in the vaccine group and none in the placebo group, but, according to FDA, there were 126 vs 111 SAEs.^17^ Pfizer’s published trial was also obscure for severe adverse events. A supplement showed that 240 patients (1.1%) had severe events on the vaccine versus 139 (0.6%) on placebo. Pfizer did not provide a P-value, but we calculated P = 2 × 10^−7^, which is highly statistically significant. The number needed to vaccinate to harm one patient severely was only 200, which was not mentioned in the study, only that “The safety profile of BNT162b2 was characterized by short-term, mild-to-moderate pain at the injection site, fatigue, and headache.”

In AstraZeneca’s trials, all of the participants in the control group received one or two doses of a meningococcal vaccine.^4^ This makes it difficult to assess the harms of the COVID-19 vaccine because the control vaccine also causes harms. The pivotal trial report noted that SAEs were less common after the COVID-19 vaccine than after the control vaccine, 79 vs 89 patients.^4^ The rate of severe adverse events was 1% but the first 14 vaccinated employees at a hospital department in Denmark became so ill after the Astra-Zeneca vaccine that all of them required a sick leave, which is by definition a severe adverse event.The discrepancy between 100% at that department and 1% in the report in *The Lancet* is so huge that we suspect AstraZeneca’s vaccine trials are unreliable. The harms were so pronounced and common that Denmark stopped using the AstraZeneca vaccine.

The mRNA vaccines can also cause severe harms. As noted above, many people were unable to perform normal daily activities after a booster with an mRNA vaccine.^76^

By far the most reliable study we identified was the systematic review that used regulatory data from the two pivotal randomised trials of the mRNA vaccines and restricted the observation period to reduce the contamination caused by offering the vaccine to patients in the placebo group.^17^ The researchers put their findings into perspective by comparing them with hospitalisations. The excess risk of SAEs of special interest was considerably larger than the reduced risk of hospitalisation, 10.1 vs 2.3 per 10,000 vaccinated people for Pfizer’s vaccine, and 15.1 vs 6.4 for Moderna’s vaccine. Even after the researchers adjusted for multiple events in the same patient in a sensitivity analysis, the risk was larger.

Serious adverse events are not directly comparable to hospitalisations. They are rarely lethal whereas a reduction in hospitalisations would be expected to reduce mortality, as some of the hospital cases were serious. On the other hand, the lower the risk of dying, the more important the serious harms are of the vaccine. These findings are therefore important for considerations about whether vaccination should be recommended for young people.

Another low-risk group involves people who have already been infected with SARS-COV-2 and recovered, and therefore have acquired natural immunity. The issue of whether to vaccinate such people is highly pertinent since most of the vaccine related harms have been attributed to over-activation of the immune system.^74^ In the only study we found, in people who had received a COVID-19 vaccine, severe harms, defined as emergency department visit or hospitalisation, occurred ten times more often if the patients had been infected earlier.^74^ Even though it was an observational study, this finding raises serious concerns about the ubiquitous recommendations to vaccinate people who have had a prior COVID-19 infection.

In the autumn of 2021, booster doses were being recommended to a largely vaccinated population,^86,87^ and in many cases mandated, worldwide. However, while it was generally accepted that the vaccines were still protective against COVID-19 hospitalisations, it was evident that protection against infection waned quickly.^88^

The data underpinning the authorisation of booster doses were based on inferior observational and immune-bridging studies, and there was great uncertainty and confusion. In December 2021, EMA recommended boosters as frequently as every three months,^89^ but in an extraordinary backflip only one month later warned that repeated boosters might weaken people’s immune responses.^90^ This has been shown to be the case for influenza vaccines. Canadian researchers, who replicated their findings in three different studies, found that people who received a seasonal influenza vaccine had an increased risk of getting infected with another strain the following year.^91^

For observational studies, the main problem is confounding. In a little known but ingenious study, a statistician used raw data from two randomised multicentre trials as the basis for observational studies that could have been carried out.^92^ He showed that the more variables that are included in a logistic regression, the further we are likely to get from the truth. He also found that comparisons may sometimes be *more* biased when the groups appear to be comparable than when they do not; that adjustment methods rarely adjust adequately for differences in case mix; and that all adjustment methods can on occasion increase systematic bias. He warned that no empirical studies have ever shown that adjustment, on average, reduces bias.^92^

Another main problem is underreporting, particularly when doctors have been reassured by authoritative messages that the vaccines are safe. In addition, there is a fear among doctors that they can be threatened with disciplinary action if they do anything that could undermine the government’s COVID-19 vaccine rollout. We have had contact with a junior doctor working in the emergency department of a major hospital who began noticing patients being rushed in with what he suspected to be serious COVID-19 vaccine injuries. His colleagues dismissed the symptoms as unrelated to the vaccine, but he felt his patients’ observations were valid. He decided to write up a report and submit it to the drug regulator but was discouraged by his head of the department as there was no protocol in place for reporting vaccine injuries. Moreover, as many doctors are stressed and overworked and do not have time to fill out the paper work, very little gets reported.

Underreporting is prevalent when the event is common in the general population, e.g. thrombosis in the elderly. Overreporting can also occur, e.g. because of increased attention related to a particular harm. In mid-March 2020, EMA warned about blood clots possibly being caused by the AstraZeneca vaccine,^80^ but the warning was downplayed so much that it was unlikely to inflate reports about vaccine injuries. EMA not only stated that “the vaccine is not associated with an increase in the overall risk of blood clots” but even that there had been *fewer* thromboembolic events than expected, both in studies before licensing and in reports after rollout of vaccination campaigns. However, EMA also noted that there had been 12 cases of cerebral venous sinus thrombosis and that only 1.4 cases was expected.

It is very unreliable to estimate expected rates. A register study found that the incidence of deep vein thrombosis in women aged 35-54 years was five times higher in USA than in Spain.^93^ The researchers also observed large variations between electronic health records and claims data sources when using the same analysis and outcome definitions. Other studies have reported a 10-fold difference in rates of transverse myelitis; a 38 times higher rate of Bell’s palsy in USA than in Italy; and a 12-fold to a 190-fold difference in rates of narcolepsy between USA and Europe.^93^

Many of the studies we reviewed were of very poor quality and published in journals that failed to identify fundamental errors. In 2021, for example, *Vaccines* (distinct from the journal *Vaccine*) published an article claiming that COVID-19 vaccines kill about as many as they save, but the authors made the basic error of assuming that all reported deaths following vaccination in pharmacovigilance data are caused by the vaccine.^94^ Tensions after its publication led to the resignation of six editors and the article was retracted a week later.^95^

In another study from the same journal, the harms were divided into mild, moderate, and severe, where mild meant lasting less than 24 hours, moderate from 24 to 72 hours, and severe more than a week.^96^ There was no category for harms lasting more than 3 and less than 8 days, and duration it not a sign of severity. A mild harm can last for weeks, and a life-threatening harm can disappear in a few minutes, e.g. an anaphylactic shock.

A systematic review used the Jadad 5-point scale for scoring the “quality” of the randomised trials.^22^ The authors claimed to have adhered to the PRISMA guidelines, but these say that “scales that numerically summarise multiple components into a single number are misleading and unhelpful.”^97^ The Jadad scale has not been recommended for the last 25 years. This review was also published in *Vaccines*.

Despite their shortcomings, we can draw some firm conclusions based on the studies we reviewed.

The adenovirus vector vaccines increase the risk of venous thrombosis and thrombocytopenia whereas we did not find reliable data in our search to suggest that COVID-19 vaccines increase the risk of arterial thrombosis. However, this area develops quickly. Our search on 4 April 2022 identified 4637 records but this number had increased by 2816 already on 2 December. A colleague notified us of a recent Israeli register study that raises concerns.^98^ It found an increase of over 25% in people aged 16-39 years in both cardiac arrest and acute coronary syndrome that was closely related to vaccination rates whereas there was no relation to COVID-19 infection rates. A Reuters Fact Check concluded that their study was misleading because it did not prove that this increase was caused by the vaccines.^99^ However, these researchers stated clearly in their paper that they had not established a causal relationship.

Infections and vaccines, e.g. against smallpox,^100^ can cause myocarditis, which is also the case for mRNA-based COVID-19 vaccines, particularly in young males. The mortality is about 1-2 per 200 cases.^28,63^

Based on biological plausibility and temporal association, inflammatory neuropathies like neuralgic amyotrophy, Bell’s palsy, and the Guillain-Barré syndrome, have been linked to other vaccines, most often to the influenza vaccine.^71^ The mounting cases of the Guillain-Barré syndrome in 1976 were closely related to the use of a swine influenza vaccine.^101^ However, serious neuropathies can be very difficult to detect. For example, it took several years before it was accepted that the influenza vaccine Pandemrix causes narcolepsy.^102,103^

When our research group analysed the clinical study reports of the HPV vaccines submitted to EMA for marketing authorisation, we showed a statistically significant increase in serious neurological adverse events.^104^ EMA denied this, and instead based its conclusion on the data provided to the agency by the manufacturers. They did not check if this reporting was accurate despite knowing that one of the companies had previously been deceptive with its HPV vaccine harms data.^11,105^

We found evidence of serious neurological harms, and a survey of 508 US patients suffering from persistent neurological symptoms after a COVID-19 vaccine showed a wide array of symptoms of which painful neuropathy/paraesthesias were the worst.^106^ Prior to the vaccination, 94% of the patients had never reacted to a vaccine. Since the symptoms are so varied, doctors tend to dismiss them and conclude that the patients suffer from a psychiatric problem. However, this is unlikely. Researchers from the US National Institutes of Health studied 23 self-referred patients who reported new neuropathic symptoms beginning within three weeks after SARS-CoV-2 vaccination (for 9 patients, after the second dose).^107^ All patients had sensory symptoms comprising severe face or limb paraesthesias, and 12 had objective evidence of small-fiber peripheral neuropathy. Autonomic testing in 12 identified 7 with reduced distal sweat production and 6 with postural orthostatic tachycardia syndrome.

Some patients have experienced similar symptoms after an HPV vaccine, which suggests autoimmunity directed against the autonomic nervous system. In a Danish study, antibodies directed against the adrenergic β-2 receptor were found in 75% of 108 patients with symptoms and in only 17% of 98 age- and sex-matched vaccinated controls (P < 0.001).^108^ Antibodies against the muscarinic M-2 receptors were found in 82% vs 16% (P < 0.001) and against either β-2 or M-2 receptors in 92% vs 19% (P < 0.001). Similar symptoms and neuroendocrine antibodies have been reported in patients suffering from long-term complications after SARS-CoV-2 infection.^108-110^

SARS-CoV-2 infection can cause transverse myelitis, with acute onset of paralysis, sensory level, and sphincter deficits due to spinal cord lesions demonstrated by imaging.^111^ The occurrence of two reported cases among 5,807 participants within two weeks after vaccination in the pivotal AstraZeneca trial,^4,111^ is an extremely high incidence considering a worldwide incidence of 0.5 cases per million after infection.^111^

SAEs have been systematically eliminated from the pivotal trials.^10^ In the Pfizer and AstraZeneca vaccine trials, participants were given digital apps to record adverse events remotely, but the apps only allowed the participants to record what the company deemed as “expected” events. If they developed thrombosis, myocarditis, Guillain-Barré Syndrome, transverse myelitis, or other serious neurological events, there was no option for them to record it on the app.

Brianne Dressen, a participant in an AstraZeneca trial, became disabled after her first injection.^10^ She is still disabled today, but there is no mention of this in the trial report in *New England Journal of Medicine*.^36^ As Dressen was concerned about the lack of reporting of her serious adverse event (and others) in the trial’s publication, she wrote to Dr Eric Rubin, editor in chief of the journal, and asked for the inaccuracies to be corrected and demanded complete reporting of the results. Rubin refused to correct the inaccurate data in his journal. The full email exchange has been made public.^10^

When Pfizer had recruited 12-15 year olds for its mRNA vaccine trial, the published data in *New England Journal of Medicine* stated that there were “no serious vaccine-related adverse events.”^46^ One of the participants, however, was 13 year old Maddie De Garay who suffered a serious adverse reaction following her second injection, which left her in a wheelchair and fed by a nasogastric tube.^10^ She was referred to hospital for a full assessment and a doctor diagnosed her with a “functional disorder.” This doctor decided she had a pre-disposition to hysteria, and she was referred to a mental health facility. Professor and psychiatrist David Healy subsequently conducted a thorough review of her medical records, including an interview with her family, and found no such history of pre-existing conditions or mental illness.

Even if data are fully reported, it is extremely difficult to find rare events in randomised trials. One would need a trial with 30,000 people in the vaccine arm to have a 95% chance of detecting a serious harm if it occurs in 1 of 10,000 cases,^112^ and one case is not enough to establish a cause-effect relationship.

A rare condition is multisystem inflammatory syndrome in children. It is much more common after SARS-CoV-2 infection than after vaccination, about 200 vs one cases per million.^113^

An important issue, which has received virtually no attention, is that vaccines have non-specific effects, which are very different for live attenuated vaccines and for non-live vaccines. Peter Aaby and co-workers have shown in several studies, that live attenuated vaccines, e.g. against measles, polio and tuberculosis, decrease mortality from other infections than the targeted one, whereas non-live vaccines increase mortality.^84,114^

Aaby’s team has also analysed the randomised trials of the COVID-19 vaccines. They found that the adenovirus vector vaccines reduced total mortality, risk ratio 0.37 (0.19 to 0.70), in contrast to the mRNA vaccines, risk ratio 1.03 (0.63 to 1.71).^79^ The difference between the two estimates was statistically significant (P = 0.03). It is a missed opportunity that nowhere in the world was the vaccine roll-out done as part of a randomised trial that could tell us if some vaccines lower mortality more than others. But now that boosters are being recommended, such trials should be performed. There is also a need for additional placebo-controlled trials, e.g. in people who have already been infected.

Another missed opportunity is that the drug regulators and other authorities have been very slow in following up signals of serious harms. In July 2021, based on medical claims data in older Americans, FDA reported detecting four potential adverse events of interest: pulmonary embolism, myocardial infarction, immune thrombocytopenia, and disseminated intravascular coagulation after Pfizer’s vaccine.^17^ FDA stated it would further investigate the findings, but the agency did not disclose its data, did not warn the doctors or the public, and 1.5 years later, had still not updated its findings.^115^

The US Centers for Disease Control and Prevention were also slow. A German researcher using VAERS and EudraVigilance comparing the disproportionality of adverse event reports for the influenza vaccine and the mRNA COVID-19 vaccines reported excess risks for four Brighton serious adverse events of special interest: cardiovascular events, coagulation events, haemorrhages, gastrointestinal events, and thromboses.^115,116^ CDC published a protocol in early 2021 for disproportionality analyses in the VAERS database,^17^ but they have not reported the results.

Given all the difficulties, obstacles with getting access to regulatory data, obfuscations, and documented underreporting, we find it likely that there are other serious harms than those uncovered so far.

Further randomised trials are needed. Authorities have recommended virtually everyone get vaccinated and receive booster doses. They do not consider that the balance between benefits and harms becomes negative in low-risk groups such as children and people who have already recovered from COVID-19 infection.^117^

## Data Availability

All data are publicly available

## Acknowledgment

We thank Peter Doshi for comments on our manuscript.

## Conflicts of interest

None.

## Funding

Institute for Scientific Freedom and a private funder requesting anonymity, a retired filmmaker with no conflicts of interests in relation to our review.

## Availability of data

The data we have published are publicly available.

## Notes

### Competing Interest Statement

The authors have declared no competing interest.

### Summary of Updates

only minor changes

